# Postpartum hormonal contraceptive use in Denmark during 1997-2021

**DOI:** 10.1101/2024.02.21.24302996

**Authors:** Søren Vinther Larsen, Anders Pretzmann Mikkelsen, Kathrine Bang Madsen, Xiaoqin Liu, Trine Munk-Olsen, Vibe Gedso Frokjaer, Øjvind Lidegaard

**Author notes:** contributed equally as senior authors (Vibe Gedso Frokjaer, Øjvind Lidegaard). **Corresponding Author:** Søren Vinther Larsen, MD, Neurobiology Research Unit, Copenhagen University Hospital – Rigshospitalet, Copenhagen, Denmark, Blegdamsvej 9, DK-2100 Copenhagen, Denmark, Mail.

## Abstract

**Introduction:** Hormonal contraception is used by over a quarter of a billion worldwide. In Denmark, 84% have used it before their first pregnancy. After pregnancy, mothers are routinely advised to consider contraception to avoid short interpregnancy intervals due to perinatal and maternal health risks. Yet, mothers are not recommended to start combined hormonal contraception within six weeks postpartum due to an increased thromboembolic risk. This study provides an overview of postpartum hormonal contraceptive use in Denmark.

**Material and methods:** This descriptive study is based on Danish national health registries on mothers who delivered during 1997-2021. The cumulative incidence of HC initiation one year after delivery is reported by calendar year and age group. Timing of initiation is reported as the median time from delivery. Hormonal contraception was categorized according to hormone type and method of administration.

**Results:** A total of 676 759 first-time and 552 142 second-time mothers were registered, with a cumulative incidence of hormonal contraceptive initiation of 41.0% (95% CI, 40.9-41.1) and 40.5% (95% CI, 40.4-40.6), respectively. From 1997 to 2021, the cumulative incidence of first-time mothers who initially used progestogen-only pills increased from 3.8% (95% CI, 3.5-4.0) to 14.4% (95% CI, 13.9-14.8) and intrauterine levonorgestrel-releasing systems from 0.1% (95% CI, 0.1-0.2) to 12.6% (95% CI, 12.3-13.0). In contrast, combined oral contraception initiation decreased from 31.3% (95% CI, 30.7-31.8) to 7.8% (95% CI, 7.5-8.2). Among first-time mothers initiating hormonal contraception, the median time of initiation decreased from 4.7 (Q1-Q3, 2.5-7.5) months during 1997-2001 to 2.5 (Q1-Q3, 2-0-4.0) months during 2017-2021. The cumulative incidence of first-time mothers using combined hormonal contraception six weeks after delivery decreased from 1.5% (95% CI, 1.5-1.6) during 2007-2011 to 0.5% (95% CI, 0.5-0.5) during 2017-2021.

**Conclusions:** Within the first year after childbirth, 41% of first– and second-time mothers initiated hormonal contraception in Denmark during 1997-2021. Throughout 1997-2021, mothers started earlier after delivery and more often used progestogen-only contraception. Few started combined hormonal contraception within 6 weeks after delivery in accordance with national guidelines. Taken together, the pattern of HC use over time reflects a change to safer contraceptive methods postpartum which minimizes thromboembolic risk.

## Introduction

Hormonal contraception (HC) is used by more than a quarter of a billion women worldwide (1). They are widely used for family planning and for treating medical conditions such as dysmenorrhea, menorrhagia, endometriosis, polycystic ovary syndrome, and acne (2–6). In the Nordic countries, 30-40% of women in the fertile age use HC (7); in 2019, this counted 39% in Denmark (8) where as many as 84% had used it before their first pregnancy (9). Little is known about HC use specifically during the postpartum period. However, family planning is equally important during this critical period to avoid unintended pregnancies with short interpregnancy intervals while at the same time avoid introducing an increased thromboembolic risk and reducing lactational capacity.

HC either contains progestin alone or in combination with estradiol or estestrol or the synthetic ethinylestradiol (10). Progestins are synthetic progestogen compounds and exist in various forms differentiated by their structural properties (11). Based on the time the progestins were introduced, they have been classified into first generation (norethisterone), second generation (levonorgestrel, norgestimate, and norgestrolmin^1^), third generation (desogestrel, gestodene, etonorgestrel), fourth generation (drospirenone), and non-classified compounds (cyproterone acetate, nomegestrol acetate, and dienogest) (8). HC also differs by the way it is administered: oral (combined oral contraception (COC) and progestogen-only pill (POP)), transdermal (patch), subdermal (implant), intramuscular (depot injections), vaginal (the vaginal ring), and intrauterine (intrauterine levonorgestrel-releasing system (IUS)).

In Denmark, postpartum contraceptive counselling is provided on a routine basis at the 8-week postpartum consultation with the general practitioner (12). These visits are part of standard antenatal care program, and here mothers are advised to consider contraception to avoid short interpregnancy intervals as this may lead to the choice of aborting the pregnancy or increased perinatal and maternal health risks (13,14). Short interpregnancy intervals occur more often when the subsequent pregnancy is unplanned (15,16). In Denmark, as many as 4% of those who delivered twice between 1994-2010 had an interpregnancy interval of less than 6 months, which was associated with higher risk of preterm birth and small for gestational age delivery (17).

On the other hand, it is not recommended to start combined hormonal contraception (CHC) too early after the delivery due to an increased risk of thromboembolic events (18,19). Therefore, the Danish Health Authority does not recommend initiating CHC before six weeks postpartum in non-breastfeeding mothers (20). Meanwhile, the World Health Organization is less restrictive recommending non-breastfeeding mothers not to use it before three weeks after delivery and for those with thromboembolic risk factors not before six weeks after delivery (21). Another reason for not considering the initiation of CHC is the potential negative impact it may have on lactational capacity (22). Taking a conservative approach, it is not recommended that mothers use it while breastfeeding in Denmark, and the World Health Organization does not recommend it until 6 months after delivery (21,23). However, we have yet no overview of whether these recommendations are followed in clinical practice. Consequently, the aim of this study was to give an overview of time trends in postpartum HC use in Denmark from 1997 through 2021, including an overview of the types used stratified by age group and parity, and of the timing of initiation after delivery.

## Methods

### Study design

This descriptive study used Danish national health registry data linked via the unique personal identification number given to Danish residents at birth or immigration. Data were provided by the Danish e-Health Authority and hosted by Statistics Denmark.

### Study population

All women giving birth to a liveborn child for the first and second time between January 1, 1997, and December 31, 2021, were identified via the Danish Civil Registration System and the Medical Birth Registry (24,25). The first and second childbirth were chosen as it covers approximately 80% of deliveries in Denmark while covering a broad maternal age range (https://www.statistikbanken.dk/fodpm).

### Hormonal contraceptives

Mothers were registered as postpartum incident users of HC on the date they filled a prescription for HC within 12 months after their delivery registered in the Danish national prescription register (26). HC types were categorized as COC with a progestin from first generation (ATC codes G03AA01, G03AA03, G03AA05, and G03AB04), second generation (ATC codes G03AA07, G03AA11, and G03AB03), third + fourth generation (ATC codes G03AA09, G03AA10, G03AA12, G03AB05, and G03AB06), COC containing either natural estrogen, dienogest, or cyproterone acetate (ATC codes G03AA14, G03AA16, G03AA18, G03AB08, and G03HB01), vaginal ring and patch (ATC codes G03AA13 and G02BB01), POP (ATC codes: G03AC01, G03AC02, G03AC03, G03AC09, and G03AC10), IUS with low (13.5 mg), medium (19.5mg), and high (52 mg) levonorgestrel content (ATC code G02BA03), implants (ATC code G03AC08), and depot injections (ATC code G03AC06). In addition, contraceptives lasting maximally three months will be referred to as short-acting contraceptives (SARC), i.e., COC, POP, Patch, vaginal ring, and depot injections, and those with longer duration as long-acting reversible contraceptives (LARC), i.e., IUS and implant.

### Statistical Analyses

Women were followed from first or second delivery until incident HC use postpartum, emigration, death, 12 months after delivery, or December 31, 2022, whichever came first. The total number and proportion of first-time mothers who initiated HC within 12 months postpartum including the distribution of types initially used are reported. Cumulative incidence curves of HC initiation after first delivery are created by 5-year calendar periods and by HC type. The Cumulative incidence is reported by calendar period and age group (defined as less than 20 years, 20-24 years, 25-29 years, 30-34 years, and 35 years and older) for first– and second-time mothers. To provide an overview of the postpartum timing of the first HC type initiated across calendar periods, we show the calendar period-specific cumulative incidence curves and median time of initiation of the first HC type initiated among the first-time mothers who initially started on the given HC type postpartum.

In addition to the overview of the first HC initiation in the postpartum period, we provide a month-by-month prevalence use of the different HC types expressed as defined daily doses per 1000 women per day for first-time mothers who delivered between 2017-2021. By such, we incorporate information about HC discontinuation and switch of type to give an overview of HC use throughout the postpartum period. To do so, consecutive prescriptions were handled as follows: If a prescription was refilled before the end of the last prescription (the duration was set to 3 years for low-dose IUS and 5 years for medium and high-dose IUS), the prescription was registered as a continuation of the last prescription. If a switch to another HC type happened less than 28 days from the end of the former HC type, the new type was registered to start in continuation of the end the former type, but if it happened more than 28 days before the end of the last prescription, the new HC type was registered to start on the day the prescription was filled, at what point the former prescription was registered to end. Postpartum discontinuation was registered when the last filled HC prescription ended within 12 months from delivery and no new prescription was filled within the duration of this prescription plus a grace period of 28 days (27).

Last, we provide an overview of the cumulative incidence of first-time mothers who started a CHC before three and six weeks and six months postpartum. All analyses were conducted using R version 4.2.2 (28).

### Ethics Statement

Approval was achieved from the Danish Data Protection Agency (journal-nr. Pactius-2020-217 and “Privacy”, 2022) and the National Data Health Board. No ethical approval or informed consent is needed for register-based studies in Denmark. (28)

## Results

Among 676 759 first-time and 552 142 second-time mothers during 1997-2021 in Denmark, the 12-months cumulative incidence of HC initiation was 41.0% (95% CI, 40.9-41.1) and 40.5% (95% CI, 40.4-40.6), respectively. This was 37.7% (95% CI, 37.5-38.0) among first-time mothers who delivered during 1997-2001, increasing to 44.4% (95% CI, 44.1-44.6) during 2006-2011, hereafter decreasing to 38.8% (95% CI, 38.5-39.0) during 2017-2021 (**Figure 1A**). The cumulative incidence of postpartum HC users was more than 50% for first-time mothers under the age of 25 years and less than 25% for first-time mothers aged 35 years and above across all calendar periods (**Table 1**).

**Figure 1.**
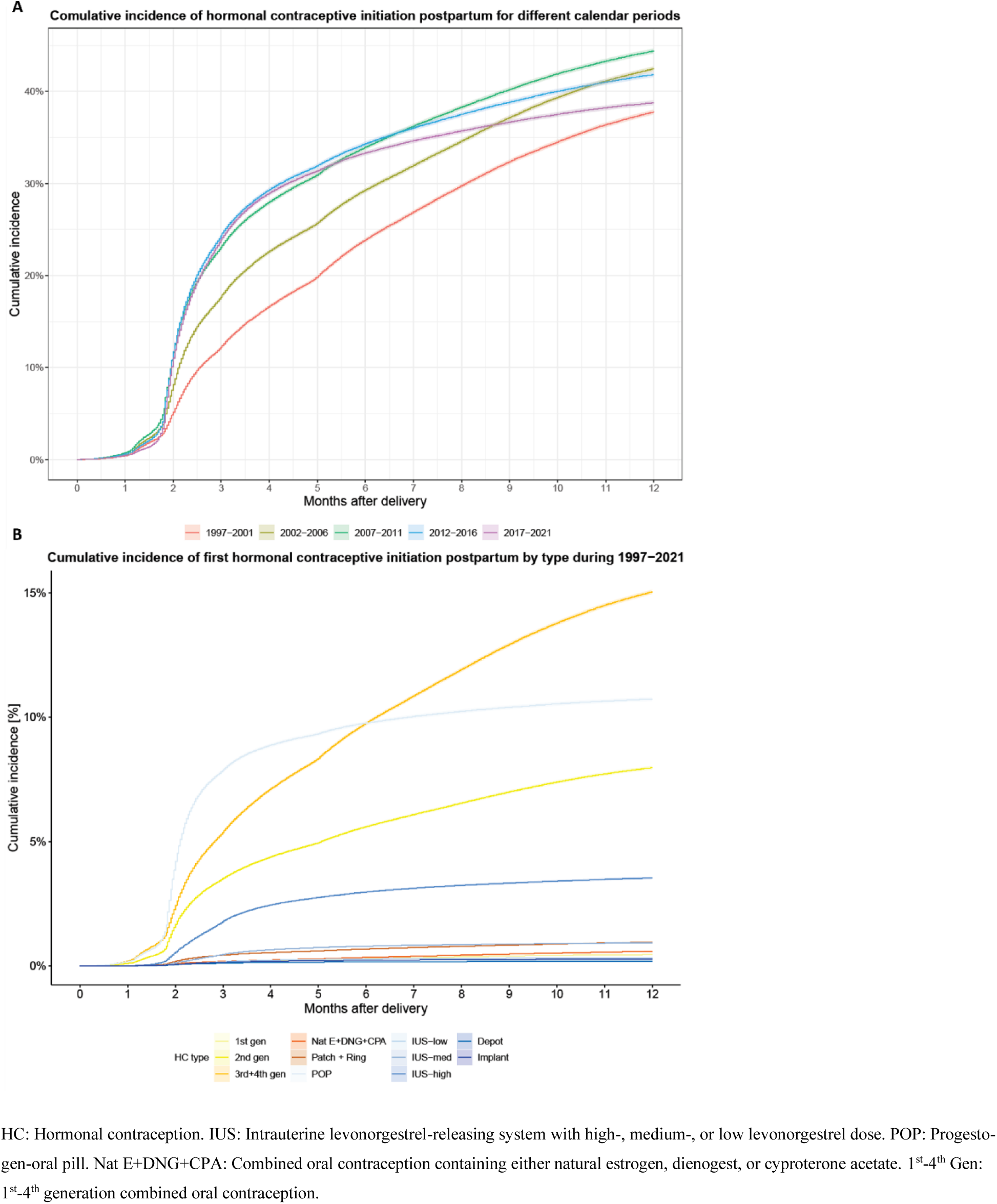
(**A**) Cumulative incidence curves of hormonal contraceptive initiation in 5-year calendar periods for first-time mothers who delivered during 1997 and 2021. **(B)** The cumulative incidence of first hormonal contraceptive initiation postpartum by hormonal contraceptive type for first-time mothers who delivered during 1997 and 2021.

**Table 1.**
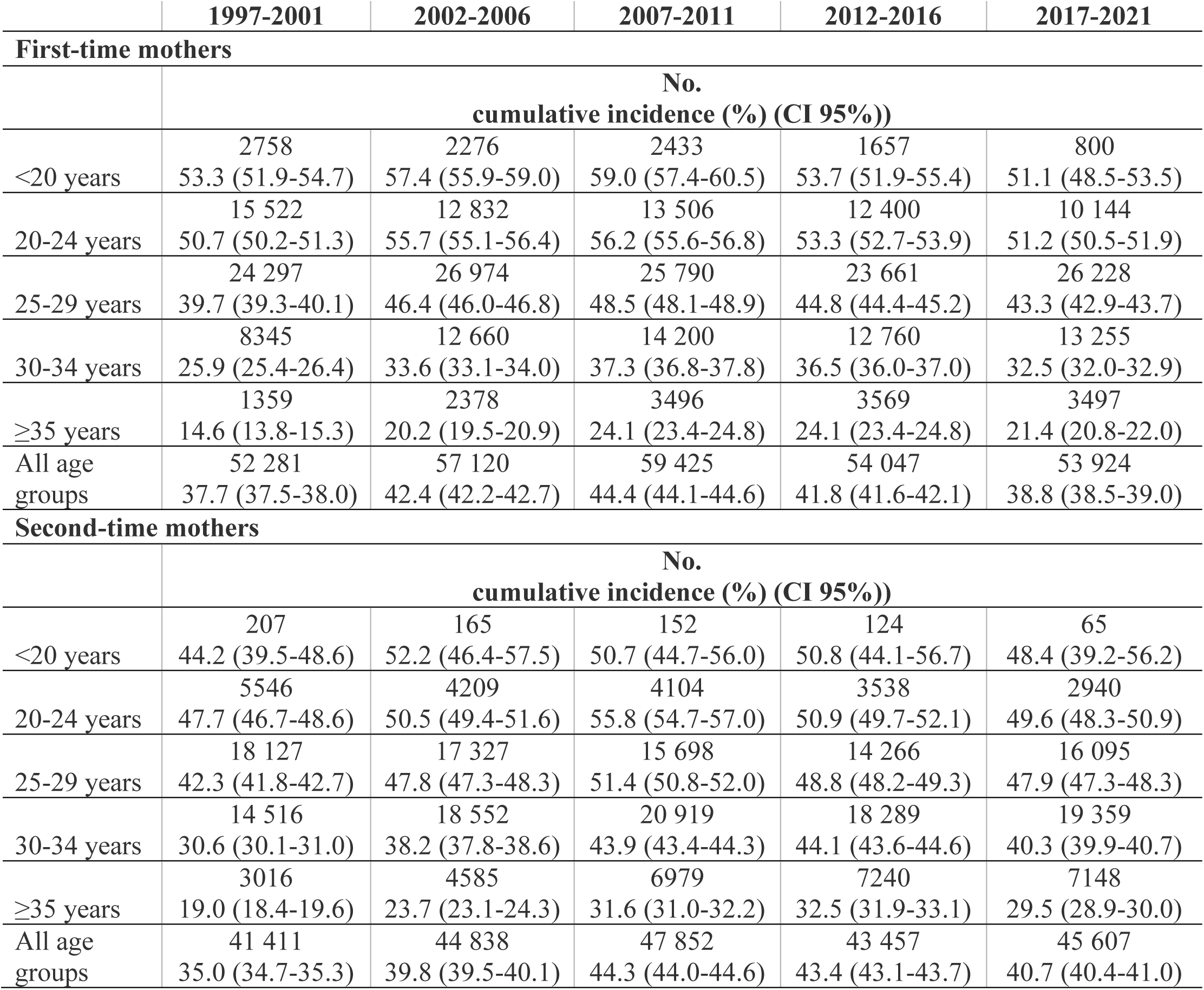
Hormonal contraceptive users within 12 months after delivery across age groups and calendar periods.

Across all calendar years, the cumulative incidence was 15.0% (95% CI, 15.0-15.1) for third and fourth generation COC, 10.7% (95% CI, 10.7-10.8) for POP, 8.0% (95% CI, 7.9-8.0) for second generation COC, 3.5% (95% CI, 3.5-3.6) for high-dose IUS, 1.0% (95% CI, 0.9-1.0) for patch and vaginal ring, 0.9% (95% CI, 0.9-1.0) for medium-dose IUS, 0.6% (95% CI, 0.6-0.6) for COC containing natural estrogen, dienogest, or cyproterone acetate, 0.5% (95% CI, 0.4-0.5) for first generation COC, 0.3% (95% CI,0.3-0.3) for low-dose IUS, 0.3% (95% CI, 0.3-0.3) for implant, and 0.2% (95% CI, 0.2-0.2) for depot injections (**Figure 1B**).

The number and proportion of first-time mothers who initiated HC postpartum are shown by calendar year in **Figure 2A** and the distribution of the HC types initially used in **Figure 2B**. The cumulative incidence of POP use as first HC after giving birth increased from 3.8% (95% CI, 3.5-4.0) to 14.4% (95% CI, 13.9-14.8) among first-time mothers who delivered in 1997 and 2021, respectively. Correspondingly, the number who initially used an IUS increased from 0.1% (95% CI, 0.1-0.2) to 12.6% (95% CI, 12.3-13.0) and, in contrast, those who initially used COC decreased from 31.2% (95% CI, 30.7-31.8) to 7.8% (95% CI, 7.5-8.2). This included a decrease in the use of 3^rd^+4^th^ generation COC, especially among first-time mothers who delivered in 2010 vs. 2012, where it decreased from 22.7% (95% CI, 22.2-23.2) to 6.8% (95% CI, 6.5-7.1). Of the first-time mothers initiating HC postpartum who delivered in 2021, 40.1% initially used POP, 35.3% IUS (14.9% a high-dose IUS, 17.6% a medium-dose IUS, and 2.8% a low-dose IUS), and 21.9% COC (**Figure 2B**).

**Figure 2.**
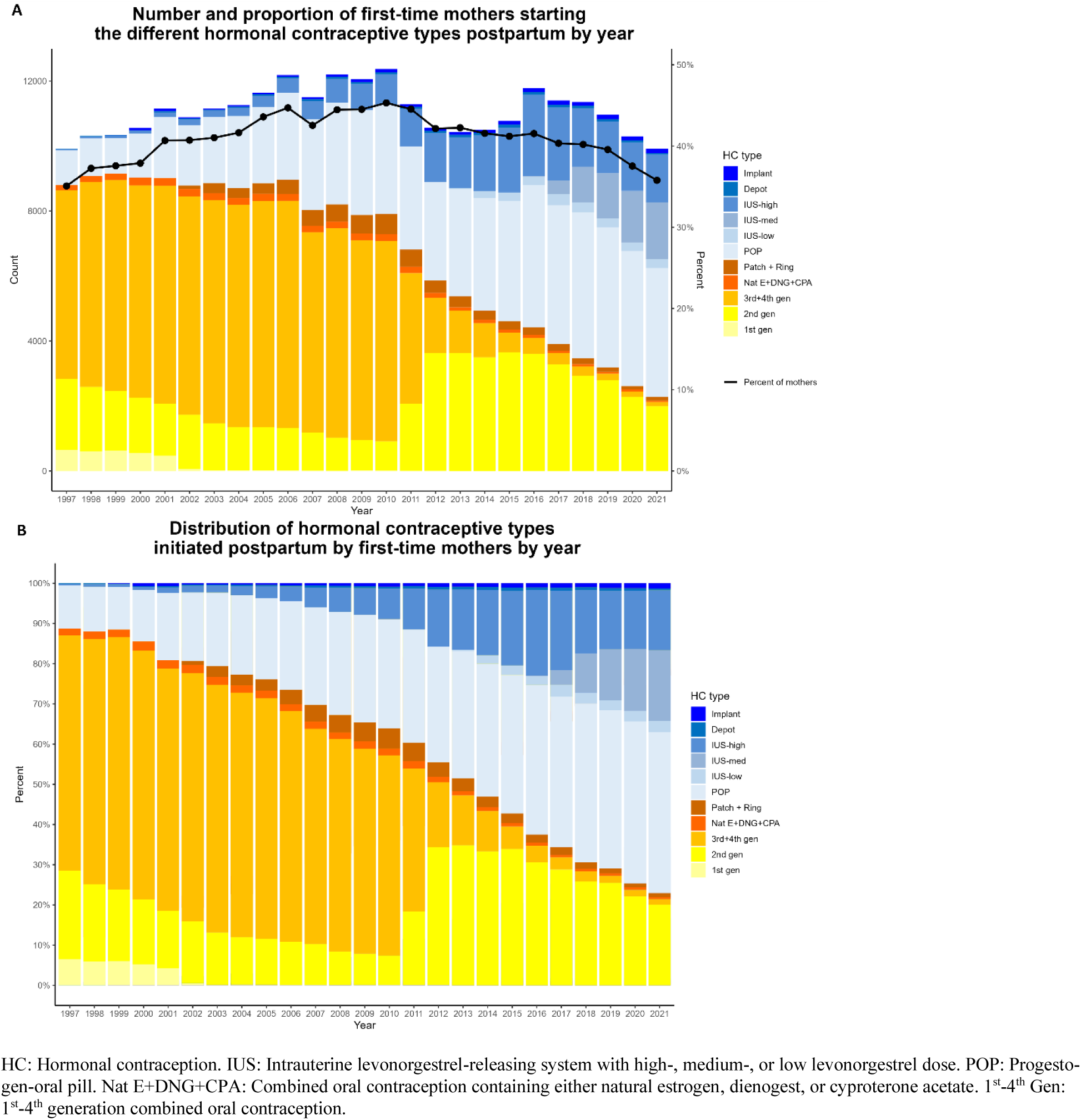
Time trends in first use of different hormonal contraceptive types within first postpartum year among first-time mothers giving birth between 1997 and 2021. (**A**) shows the proportion of first-time mothers who initiated hormonal contraception postpartum and the total number of first-time mothers who initiated the different hormonal contraceptive types. (**B**) shows the distribution of the different hormonal contraceptive types among users of hormonal contraception postpartum.

The proportion of first– and second-time mothers during 2017-2021 who initiated HC postpartum are shown in **Figure 3A** and **Figure 3C**. Of the first-time mothers initiating HC postpartum who delivered between 2017-2021, 47.6% of the adolescent mothers (<20 years of age) initially used CHC (**Figure 3B**). In contrast, this was 18.6% in those 35 years or older. Across all age groups, 31.4% initially used an LARC (**Figure 3B**). In comparison, this was 53.4% among second-time mothers (**Figure 3D**).

**Figure 3.**
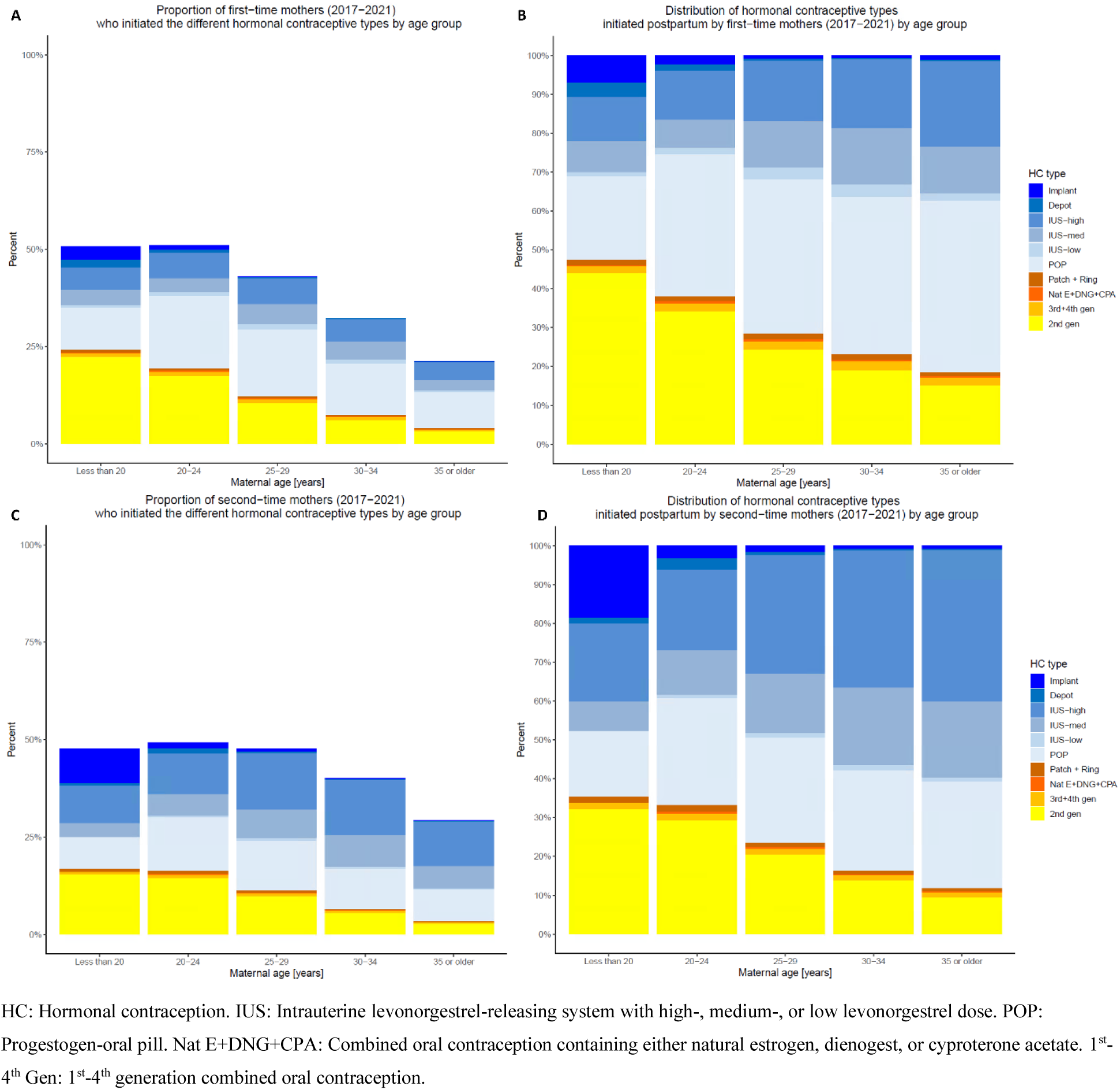
Among mothers in the different age groups who delivered for the first– or second time between 2017-2021, (**A**) shows the proportion of first-time mothers who initially started on the different hormonal contraceptive types in the postpartum period, (**B**) shows the distribution of the different hormonal contraceptive types initiated by the first-time mothers in the postpartum period, (**C**) shows the proportion of second-time mothers who initially started on the different hormonal contraceptive types in the postpartum period, (**D**) shows the distribution of the different hormonal contraceptive types initiated by the second-time mothers in the postpartum period.

First-time mothers during 2017-2021 initiated HC earlier after delivery than in the preceding calendar periods (**Figure 4A-E**), with a median time of initiation after delivery of 2.5 (Q1-Q3, 2.0-4.0) months, which was more than two months earlier than among first-time mothers between 1997-2001 where the median time was 4.7 (Q1-Q3. 2.5-7.5) months (**Table 2**). Initiation of POC happened earlier after delivery than initiation of the CHC, but this difference attenuated across the calendar periods (**Figure 4A-E**); The median time of POP initiation decreased from 2.3 (Q1-Q3, 1.9-3.5) months to 2.2 (Q1-Q3, 1.9-3.1) months among first-time mothers who delivered between 1997-2001 and 2017-2021, respectively. The corresponding median times for second generation COC were 5.3 (Q1-Q3, 2.9-8.0) months and 2.7 (Q1-Q3, 2.0-5.3) months (**Table 2**).

**Figure 4.**
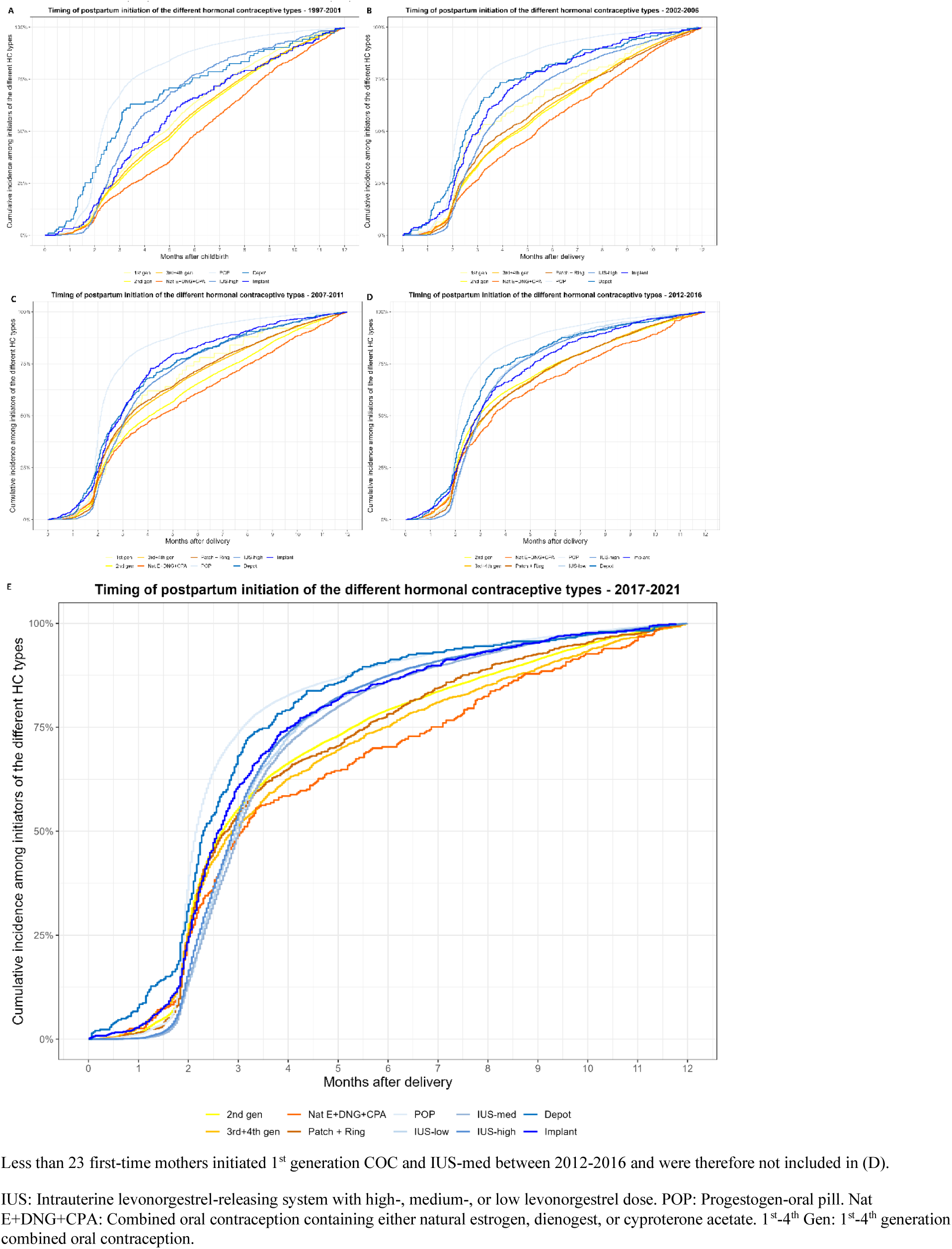
Hormonal contraceptive type-specific cumulative incidence curves among initiators of each type in first-time mothers who delivered between (**A**) 1997-2001, (**B**) 2002-2006, (**C**) 2007-2011, (**D**) 2012-2016, and (**E**) 2017-2021.

**Table 2.** Median time from delivery to hormonal contraceptive initiation.

|  | 1997-2001 | 2002-2006 | 2007-2011 | 2012-2016 | 2017-2021 |
| --- | --- | --- | --- | --- | --- |
|  | Months (Q1-Q3) |  |  |  |  |
| Combined hormonal contraceptives |  |  |  |  |  |
| 1<sup>st</sup> gen | 4.8 (2.7-7.3) | 3.1 (2.1-6.9) | 3.3 (2.3-5.8) | N/A | N/A |
| 2<sup>nd</sup> gen | 5.3 (2.9-8.0) | 4.7 (2.5-7.7) | 4.1 (2.2-7.5) | 2.9 (2.0-6.0) | 2.7 (2.0-5.3) |
| 3<sup>rd</sup>+4<sup>th</sup> gen | 5.1 (2.8-7.8) | 4.5 (2.5-7.6) | 3.4 (2.1-6.6) | 3.2 (2.0-6.2) | 2.9 (3.0-5.9) |
| Nat E+DNG+CPA | 6.2 (3.5-8.7) | 5.4 (2.8-8.4) | 4.5 (2.3-8.1) | 3.5 (2.1-7.0) | 3.1 (2.0-6.9) |
| Patch + Ring | N/A | 4.1 (2.3-7.5) | 3.3 (2.1-6.5) | 3.2 (2.1-6.1) | 2.8 (2.0-5.6) |
| Progestogen-only contraceptives |  |  |  |  |  |
| POP | 2.3 (1.9-3.5) | 2.2 (1.9-3.1) | 2.1 (1.9-3.0) | 2.1 (1.9-3.1) | 2.2 (1.9-3.1) |
| IUD-low | N/A | N/A | N/A | 3.0 (2.3-4.5) | 3.0 (2.3-4.2) |
| IUD-med | N/A | N/A | N/A | N/A | 3.0 (2.3-4.0) |
| IUD-High | 3.5 (2.5-5.7) | 3.4 (2.4-5.9) | 3.2 (2.3-5.3) | 3.0 (2.2-4.5) | 2.9 (2.2-4.1) |
| Depot | 2.9 (1.7-5.9) | 2.5 (1.9-4.5) | 2.9 (1.9-5.1) | 2.6 (1.9-4.2) | 2.3 (1.9-3.6) |
| Implant | 4.6 (2.7-7.3) | 3.0 (2.1-4.9) | 2.9 (2.0-4.5) | 2.9 (2.1-5.1) | 2.6 (2.0-4.0) |
| All types | 4.7 (2.5-7.5) | 3.6 (2.2-7.0) | 2.9 (2.0-5.7) | 2.6 (2.0-4.8) | 2.5 (2.0-4.0) |

Less than 23 first-time mothers initiated 1<sup>st</sup> generation COC and IUS-med between 2012-2016 so therefore the median times of initiation of these were not included.
IUS: Intrauterine levonorgestrel-releasing system with high-, medium-, or low levonorgestrel dose. POP: Progestogen-oral pill. Nat E+DNG+CPA: Combined oral contraception containing either natural estrogen, dienogest, or cypoterone acetate. 1<sup>st</sup>-4<sup>th</sup> Gen: 1<sup>st</sup>-4<sup>th</sup> generation combined oral contraception.

The postpartum month-by-month prevalence of HC use represented by defined daily doses per 1000 mothers per day for first-time deliveries during 2017-2021 is shown in **Figure 5A**. The prevalence increased from the first to the fourth month postpartum, whereafter it reached a relatively stable level at around 26-29% for the rest of the postpartum period. POP was more frequently used in the early phase after delivery, reaching 50.8% of all prescribed HC in the third month, hereafter it declined to 25.5% in the twelfth month after delivery (**Figure 5B**). In contrast, IUS use increased from 21.2% to 41.6%, and COC steadily increased from 25.1% to 29.8% from the third to the twelfth month postpartum. Among those who initially used POP, 39.2% discontinued and 16.8% switched to another type (mainly second generation COC, which counted 13.0%) within the postpartum period (**Figure S1**). Notably, 24.4% only filled one POP prescription, which was 17.6% for COC, 26.1% for patch and vaginal ring, and 23.2% for depot injection (it is uncertain for LARC as the duration of these extends beyond the postpartum period and little information exist about removal of these). During the first two months after delivery, very few mothers used contraception, but of those who did, a relatively larger fraction used CHC (44.1% in the first and 33.7% in the second month) than in the following months postpartum.

**Figure 5.**
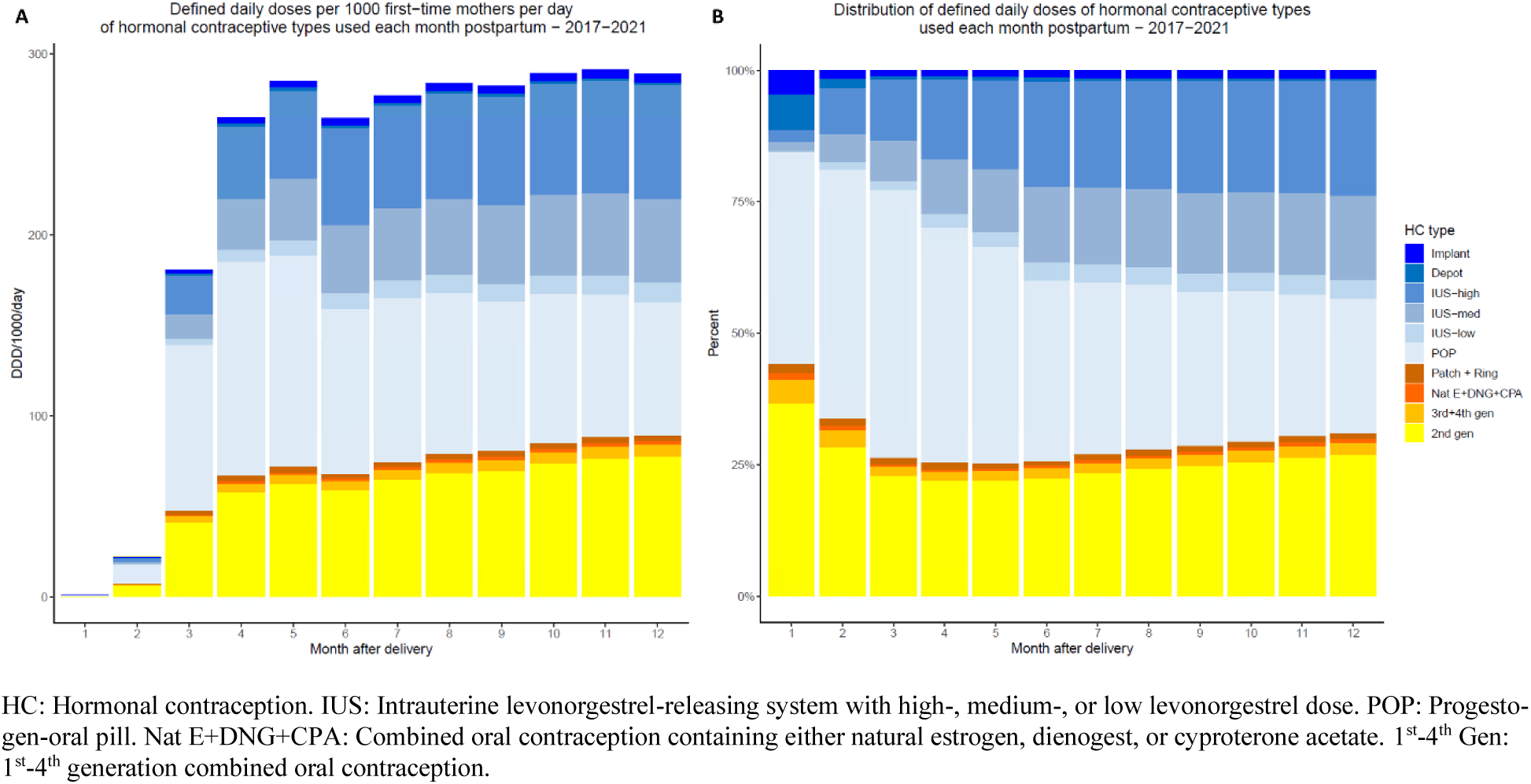
(**A**) The month-by-month defined daily doses per 1000 mothers per day after first-time deliveries between 2017-2021. (**B**) The month-by-month distribution of defined daily doses between the different hormonal contraceptive types used by first-time mothers who delivered between 2017-2021.

Among first-time mothers who delivered between 1997-2001, 1.1% (95% CI, 1.0-1.1)) started a CHC within six weeks postpartum. This increased to 1.5% (95% CI, 1.5-1.6) among those who delivered between 2007-2012 and decreased again to 0.5% (95% CI, 0.5-0.5) for those delivering between 2017-2021 (**Table 3**). Of all the first-time mothers who initiated a HC postpartum, this corresponded to 2.8%, 3.5% and 1.3% for the respective calendar periods. For the corresponding calendar periods, 19.6% (95% CI, 19.4-19.9), 21.4% (95% CI, 21.2-21.6), and 9.5% (95% CI, 9.3-9.6) of first-time mothers started CHC within six months postpartum.

**Table 3.**
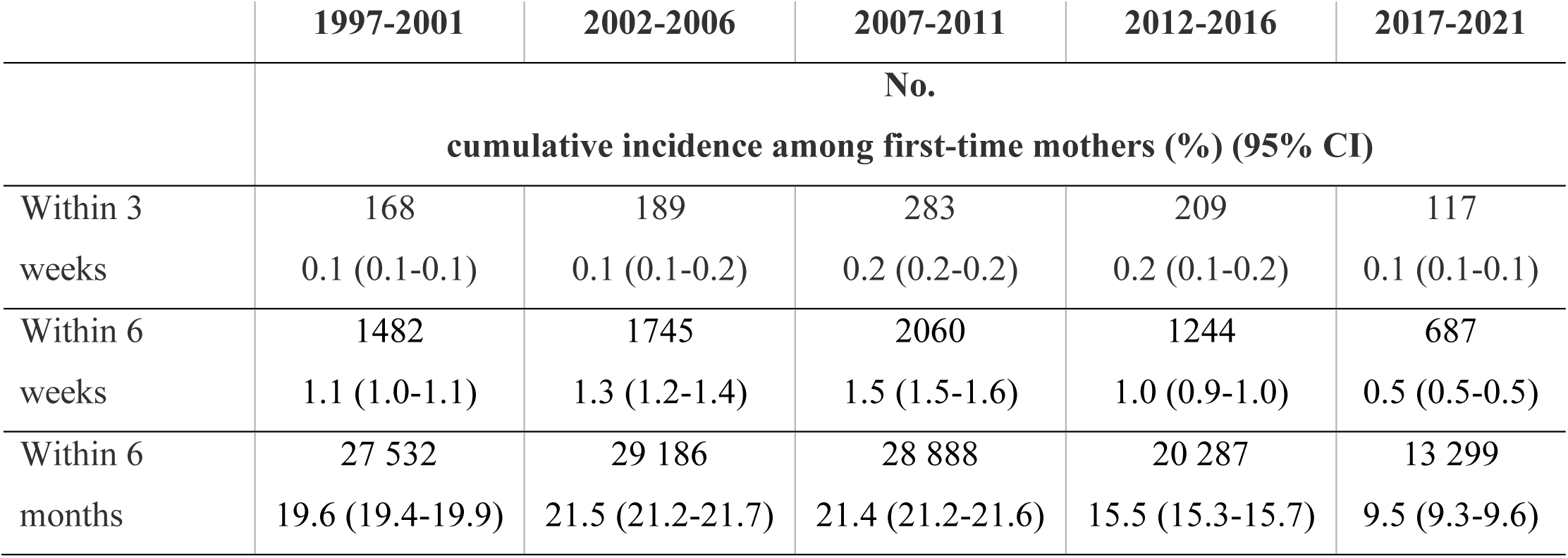
First-time mothers starting a combined hormonal contraceptive within 3 weeks, 6.

|  | **1997-2001** | **2002-2006** | **2007-2011** | **2012-2016** | **2017-2021** |
| --- | --- | --- | --- | --- | --- |
|  | **No.**<br>**cumulative incidence among first-time mothers (%) (95% CI)** |  |  |  |  |
| Within 3 weeks | 168<br>0.1 (0.1-0.1) | 189<br>0.1 (0.1-0.2) | 283<br>0.2 (0.2-0.2) | 209<br>0.2 (0.1-0.2) | 117<br>0.1 (0.1-0.1) |
| Within 6 weeks | 1482<br>1.1 (1.0-1.1) | 1745<br>1.3 (1.2-1.4) | 2060<br>1.5 (1.5-1.6) | 1244<br>1.0 (0.9-1.0) | 687<br>0.5 (0.5-0.5) |
| Within 6 months | 27 532<br>19.6 (19.4-19.9) | 29 186<br>21.5 (21.2-21.7) | 28 888<br>21.4 (21.2-21.6) | 20 287<br>15.5 (15.3-15.7) | 13 299<br>9.5 (9.3-9.6) |

## Discussion

This study shows that the prevalence of postpartum HC initiation in Denmark increased from 38% of first-time mothers between 1997-2001 to 44% between 2007-2011 and hereafter decreased to 39% between 2017-2021. From 1997 to 2021, the first HC type used shifted from COC to POC, making up more than two-thirds of HC types initially used by first-time mothers giving birth in 2021. Between 1997 and 2021, the median time until HC initiation after delivery among first-time mothers was shortened by more than 2 months. POC was generally initiated earlier after delivery than CHC, a difference that, however, attenuated over the years. Since many first-time mothers during 2017-2021 discontinued HC use already during the postpartum period, the prevalence of HC use 12 months postpartum did not reach more than 29%.

The cumulative incidence of HC initiation within 12 months postpartum in Denmark was comparable to in the United States, where 41.7% of women in the US military healthcare system filled a HC prescription within 12 month postpartum (29). In contrast, the cumulative incidence of postpartum HC initiation was higher in Denmark compared to low– and middle-income countries in Asia and Africa, where modern contraceptive initiation also reached 41% one year postpartum, however, this included sterilization and barrier methods too (30). In these countries, depot injection was used with high frequency in the postpartum period, which was not the case in Denmark.

In comparison to outside the postpartum period in Denmark in 2019 (8), the prevalence of HC use was about 10% lower in the postpartum period among mothers giving birth between 2017-2021 (not counting the first three months postpartum), and POP was more often used by HC users counting between 26-51% dependent on the postpartum month vs. 9% among HC users outside the postpartum period. This was opposite to COC use, which was used by about 50% of HC users outside the postpartum period vs. 25-30% in the postpartum period. Similar to outside the postpartum period, HC use was more prevalent in women younger than 25 years compared older age groups (8).

About one-third of first-time mothers used LARC similar to outside the postpartum period; however, it was more often used in second-time mothers, with more than 50% initiating LARC in women 35 years or older. This possibly reflects that more first-time mothers than second-time mothers wished to have another child within a relatively short period of time after the first, for whom LARC, which last three to six years, were less appropriate.

A relatively large fraction of 18-26% of first-time mothers who initially used a SARC discontinued after only having filled one prescription. This may be explained by multiple reasons: 1) They may have initiated at a later timepoint than they filled the prescription and thus may be misclassified as discontinuers, 2) they only used HC for a short period of time as they intended to become pregnant again, or 3) they stopped due to side effects or other HC-related reasons. Previous reports from the US indicates that as many as 20% of unintended pregnancies can be attributed to discontinuation of oral contraceptives where one of the reasons for discontinuation is side effects (31). Since as many as 40% of pregnancies among those who became pregnant again within 9 months after a former delivery have been reported as unintended (15), it would be relevant for future research to clarify if early HC discontinuation due to side effects increases the risk of unintended pregnancies with short interpregnancy intervals and if these mothers could benefit from follow-up contraceptive counselling on alternative reliable contraceptive methods to avoid unintended pregnancies.

Mothers initiated POC earlier after delivery than CHC, and POP was the preferred choice in the early postpartum phase, even though the use of POP attenuated across the postpartum period, partly due to a switch to second generation COC. This is in line with guidelines from the Danish Health Authority, which recommend that POC is preferred over combined contraceptives in breastfeeding mothers as estrogen-containing contraceptives may putatively reduce lactational capacity (23). Correspondingly, the World Health Organization recommends breastfeeding mothers not to use them within six months after delivery (21). In Denmark, 9.5% of first-time mothers who delivered between 2017-2021 filled a prescription of CHC within six months after delivery, corresponding to 24.7% of all HC initiators in the postpartum period. The relatively large fraction of CHC users during the first two months postpartum may be those who did not breastfeed or who, for other reasons, may have been in need of CHC. The proportion of mothers breastfeeding at six months postpartum has been estimated to be approximately 60-70% in Denmark (32). We do not know from this register data how many of the breastfeeding mothers used CHC. Also, we do not know whether the earlier onset after delivery of CHC use can be attributed to non-breastfeeding mothers; however, the large reduction in the prevalence across the 25-year period may indicate increasing adherence to existing guidelines for clinical practice.

The guidelines are, however, based on evidence of moderate to low quality (21,22), and as CHC may be a better solution for some women than the POC, more research is needed to support or update current recommendations.

The finding that mothers of higher maternal age, to a larger degree, preferred POC over CHC than younger mothers likely reflects the avoidance of estrogen-containing contraceptives due to the increasing risk of thromboembolic events with increasing age (33). In contrast to the Danish Health Authority, which does not recommend CHC use within six weeks after delivery due to the increased thromboembolic risk (20), the World Health Organization guidelines state that no use is recommended within three weeks after delivery and only for mothers with thromboembolic excess risk not within six weeks after delivery (21). Very few mothers initiate combined contraceptives within 3 weeks after delivery in Denmark, of mothers delivering between 2017-2021 this counted 0.1%. The number of mothers initiating within six weeks increased from 1.1% in 1997-2001 to 1.5% in 2007-2011, but decreased hereafter to 0.5% in 2017-2021, which can be attributed to the very large change in the prescription patterns of third– and fourth generation COC after 2010, where evidence emerged showing a higher thromboembolic risk related to these compared to first– and second generation COC (34). The very low prevalence of CHC use within the first six weeks after delivery shows strong adherence to clinical guidelines aimed at minimizing thromboembolic risk among postpartum women in Denmark.

A limitation should be mentioned when interpreting the results of this study, which is the assumption that mothers started HC on the day they filled a prescription as there may have been a delay or they may never have initiated it, which is highlighted by the relatively large fraction who discontinued after only having filled one prescription of an SARC.

## Conclusions

The prevalence of HC initiation in the postpartum period was fairly stable at approximately 40% for mothers giving birth between 1997 and 2021, and approximately 30% were current users one year postpartum. During the 26-year period, the time from delivery to start of HC use decreased by two months, mainly due to a shift in preference from COC to POP and IUS and because COC was initiated sooner after delivery. Among first-time mothers between 2017-2021, more than 50% initiated POC and this was even more pronounced in the older age groups. The timing of initiating CHC adhered to national guidelines, with only 0.5% starting within six weeks after delivery in 2017-2021. The shift from CHC to POC and the low prevalence of CHC use within six weeks postpartum indicates a change to safer contraceptive methods in the postpartum period in Denmark from 1997 to 2021 in terms of minimizing the risk for thromboembolic events and promoting lactation.

## Data Availability

Danish national health register data cannot be
distributed, but access to the data can be granted by the appropriate authorities

## Author Contributions

Conceptualization and methodology: All authors

*Project administration, data curation, investigation, formal analysis, validation, and visualization:* Larsen

*Resources and software:* Lidegaard

*Supervision:* Mikkelsen, Liu, Bang-Madsen, Lidegaard, Frokjaer.

*Writing – original draft:* Larsen

*Writing – review & editing*: All authors

*Funding acquisition.* Frokjaer

## Data accessibility statement

Danish national health register data cannot be distributed, but access to the data can be granted by the appropriate authorities.

Conflict of interest

VGF has received honorarium as a speaker for Lundbeck Pharma A/S, Janssen-Cilag A/S and Gedeon-Richter A/S. Juliane Marie Center has received research funding from Exeltis. KBM has received speaker’s fee from Medice Nordic within the last three years. TMO has received a speaker honorarium from Lundbeck Pharma A/S. The rest of the authors report no conflicts of interest.

## Funding information

The study was funded by The Independent Research Fund Denmark (grant identifier: 0134-00278B and 7025-00111B). The funder had no role in the design and conduct of the study; collection, management, analysis, and interpretation of the data; preparation, review, or approval of the manuscript; and decision to submit the manuscript for publication.

## Supplementary material

**Figure S1.**
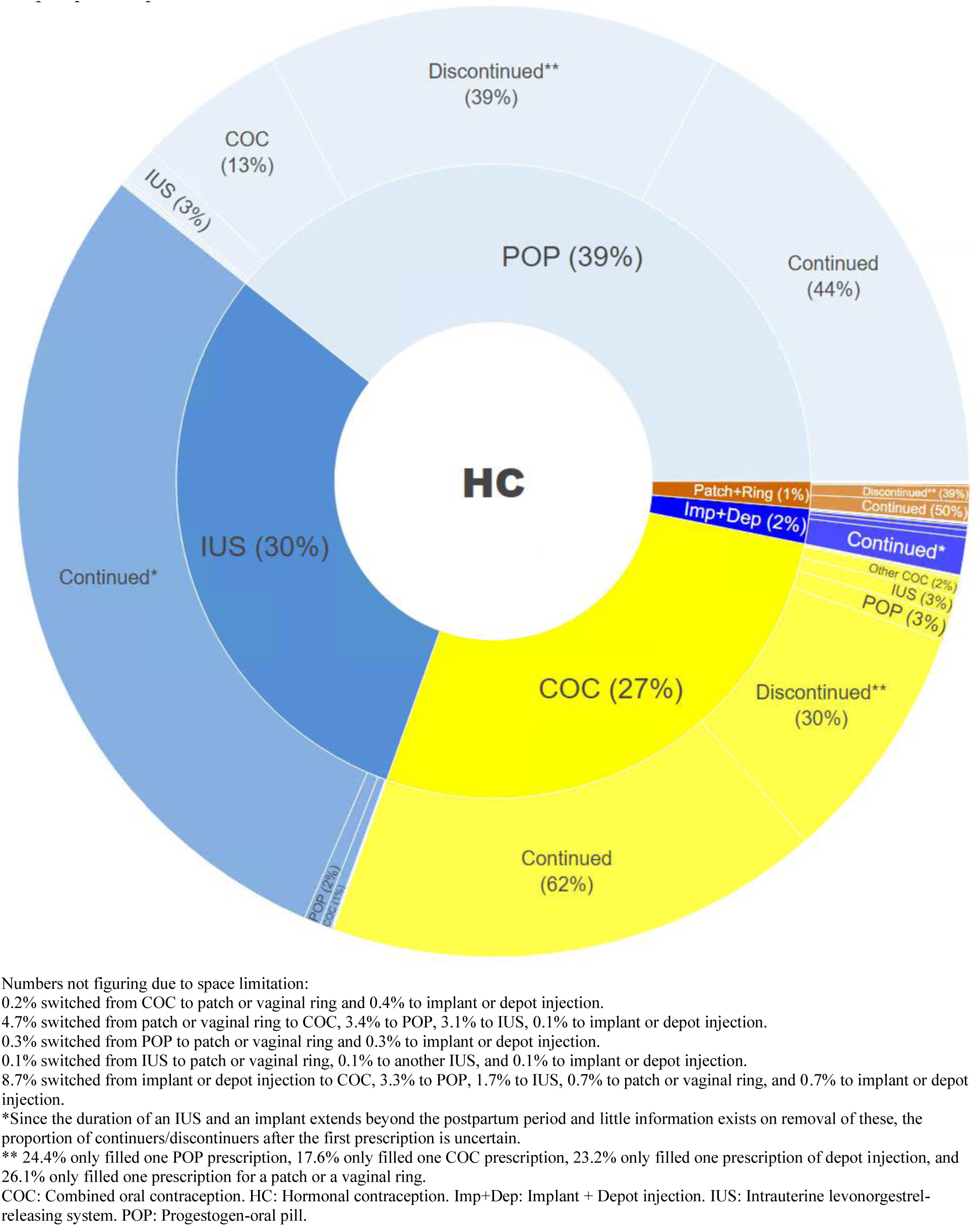
Sunburst plot showing the distribution of the hormonal contraceptive types first initiated postpartum by first-time mothers delivering during 2017-2021 and the fraction who continued, discontinued or switched (only first switch is shown) to another hormonal contraceptive type throughout the postpartum period.

## Footnotes

1 Norgestimate and norgestrolmin are often classified as a third generation progestin in the literature, but in Denmark they are traditionally classified as a second generation due to their structural similarity to levonorgestrel (20)

## Notes

### Author Declarations

This is a register study. No ethical approval or informed consent are required for register-based studies in Denmark. Danish e-Health Authority gave approval for this work. The Regional Danish Data Health Board, Privacy, gave approval for his study, journal-nr. Pactius-2020-217

